# Binary Classification of Consciousness Using Cerebral Blood Flow and EEG Features

**DOI:** 10.64898/2025.12.09.25341792

**Authors:** Farzad Azizi Zade, Irfaan Dar, Brandon Foreman, Ulas Sunar

**Affiliations:** Dept of Mechanical Engineering, Ferdowsi University of Mashhad, Mashhad, Iran; Dept of Biomedical Engineering, Stony Brook University, Stony Brook, NY, USA; Dept of Neurology & Rehabilitation Medicine, University of Cincinnati, Cincinnati, OH, USA; Dept of Electrical and Computer Engineering, Stony Brook University, Stony Brook, NY, USA

**Keywords:** disorders of consciousness, comatose, EEG, cerebral blood flow, low-frequency oscillations, machine learning, bedside monitoring, neurocritical care

## Abstract

**Background:** Assessing consciousness at the bedside in the neurocritical care unit is complicated by sedation and other treatment effects. While EEG is commonly used, it offers a limited view of the neurovascular unit. We evaluated whether combining cerebral blood flow (CBF) features with EEG improves binary classification of consciousness in patients with severe brain injury.

**Methods:** We retrospectively analyzed 35 adults who underwent multimodal neuromonitoring. Signals were segmented into 30-min windows after each probe recalibration. We used parameters including CBF low-frequency bands (Band IV 0.027-0.073 Hz, Band V 0.01-0.027 Hz, and Band All 0-0.5 Hz) and EEG band powers (delta-beta), alpha-delta ratio (ADR), alpha/(delta+theta) (ADTR), total power. A random forest (RF) model trained using K-fold cross-validation achieved optimal classification. Highly correlated features (r > 0.8) were excluded from simultaneous use. Performance was summarized with ROC-AUC and accuracy, with confusion matrices shown for the best combinations.

**Results:** Multimodal feature combinations significantly improved classification compared to EEG features alone. The best-performing combination (EEG ADR, total EEG power, and CBF Band V) achieved a ROC-AUC of 0.86 and an accuracy of 82%, representing up to 69% improvement over EEG-only models. This model also performed well on noninvasive optical blood flow data.

**Conclusions:** Combining EEG and CBF metrics, particularly low-frequency oscillations in perfusion fluctuations, enhances classification of consciousness in critically ill patients and may support future bedside tools for real-time neurovascular monitoring and in decision making about treatment and rehabilitation.

## 1 Introduction

Accurate assessment of consciousness in critically ill patients remains a major challenge in neurocritical care [1,2]. Traditional tools such as the Glasgow Coma Scale (GCS) and Coma Recovery Scale-Revised (CRS-R) are limited by sedation, paralysis, and examiner variability, leading to misdiagnosis in up to 40% of patients with covert cognitive function [3,4]. This has led to growing interest in quantitative neuromonitoring strategies that capture neurophysiological signatures of brain activity independent of behavioral output.

Electroencephalography (EEG) is widely used for bedside monitoring of brain activity due to its high temporal resolution and accessibility [5–9]. Recent studies using EEG to detect task-related or spontaneous neural responses have demonstrated that a subset of behaviorally unresponsive patients retain covert signs of preserved cortical function [4,5,10]. However, EEG provides limited insight into cerebral perfusion and is prone to artifact, low spatial resolution, and reduced sensitivity to deep or diffuse brain injury. Cerebral blood flow (CBF), by contrast, reflects the metabolic and vascular status of the brain and may capture slow, low-frequency changes related to consciousness transitions [6,11–15]. Prior studies have shown that low-frequency oscillations (LFOs) in both EEG and perfusion carry diagnostic and prognostic information in comatose patients [16–20].

Functional near-infrared spectroscopy (fNIRS) has shown promise for detecting residual brain activity using resting-state and task-based paradigms [21–26]. A recent study combining resting-state fNIRS and EEG demonstrated high classification accuracy for consciousness assessment based on low-frequency oscillations and neurovascular coherence [22]. However,

Commercialized fNIRS can measure cerebral oxygen saturation in humans but cannot provide a direct measure of cerebral blood flow (CBF) continuously [27,28]. Furthermore, oxygen saturation alone may not be sensitive enough to detect secondary brain injury hours after the insult as oxygen consumption and delivery reach equilibrium after acute transients [27,29,30]. A more recently developed diffuse optical technique, diffuse correlation spectroscopy (DCS), probes speckle fluctuations related to blood flow [28,31,32]. It is ideal for high-risk populations, as it can be used as a portable, non-invasive, continuous bedside monitor for CBF [28,33,34]. DCS blood flow contrast was shown to have a superior ability to detect response to different physiologic stimuli, as compared to fNIRS-quantified hemoglobin concentrations [35]. Additionally, we have recently shown that DCS can elucidate resting state functional connectivity (RSFC)[36–39]. DCS has been used for continuous noninvasive measurement of CBF in animals and humans [28,33,34,40–43] and very recently in NeuroICU settings [44–48].

In this study, we retrospectively analyzed multimodal EEG and cerebral blood flow (CBF) data from 35 patients in a neuroscience ICU who underwent invasive thermal diffusion flowmetry (TDF) as part of standard multimodality neuromonitoring. Patients were stratified into binary consciousness groups based on documented command following. We extracted physiologically meaningful features from EEG and invasive CBF signals, including alpha-delta ratio (ADR), total EEG power, and blood flow low-frequency oscillations (LFOs), and trained multiple machine learning classifiers to predict binary consciousness state.

To evaluate feasibility of applying the same framework to noninvasive monitoring, we additionally tested the trained models on two patients monitored with DCS. Our aim was to determine whether combining EEG and perfusion metrics improves classification accuracy compared to EEG alone, and whether noninvasive CBF features can reproduce the trends observed with invasive monitoring.

## 2 Method

### 2.1 Study overview

Figure 1 illustrates the comprehensive pipeline for this study. The process initiates with data collection, followed by segmentation and labeling. To tackle data imbalance and ensure reliability, the dataset undergoes weighing and normalization. Subsequently, it is partitioned into K-Folds (K=5) to facilitate optimization, training, and cross-validation of models.

**Figure 1.**
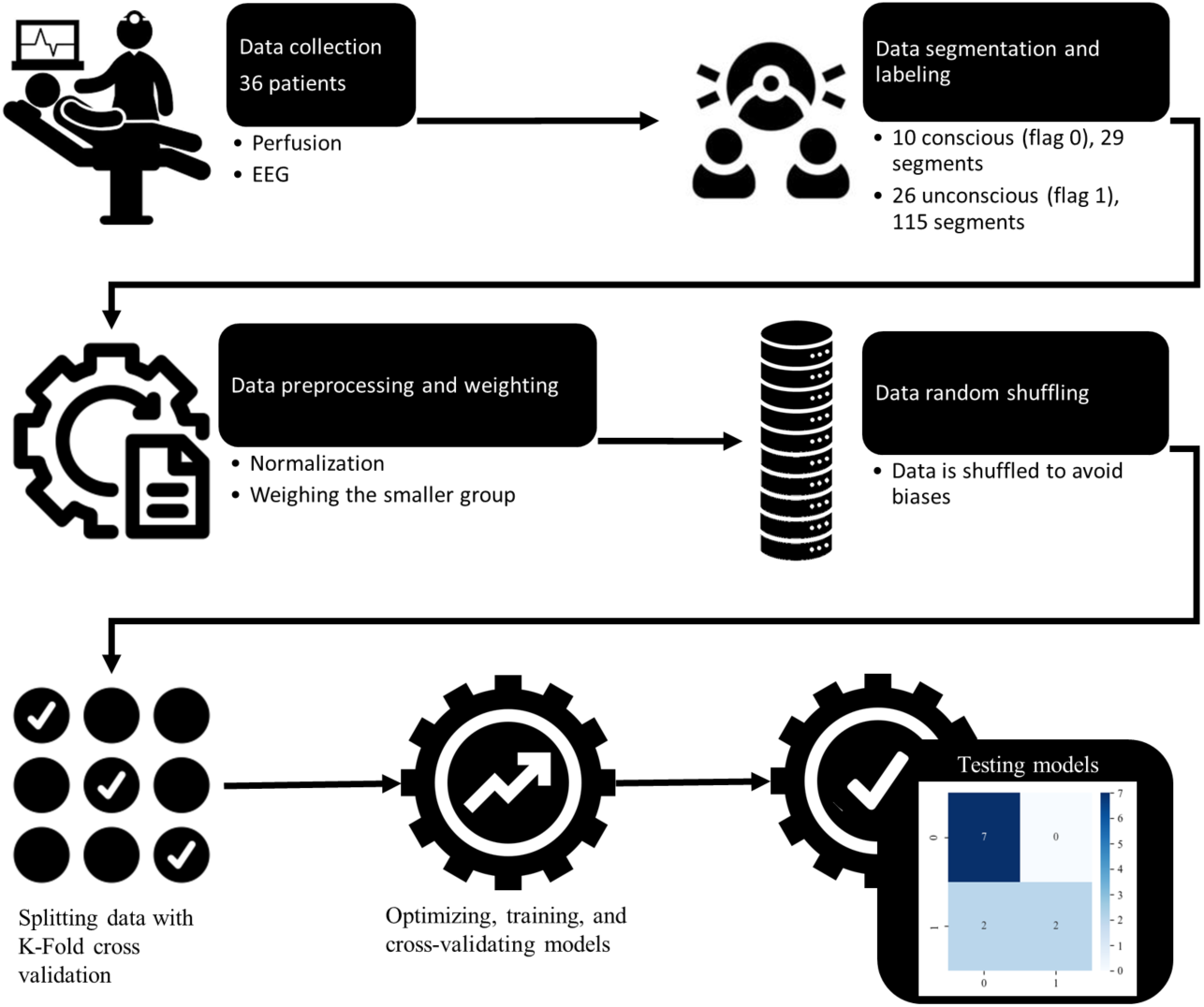
Study pipeline.

### 2.2 Patients and data collection

Adult patients admitted to the University of Cincinnati neurosciences intensive care unit (NSICU) undergoing multimodal neuromonitoring (MNM), which included invasive monitoring of regional cerebral blood flow and EEG were analyzed All patients were comatose at the beginning of MNM monitoring, and MNM was performed as part of clinical care at the discretion of the clinical care team. Research was approved with waiver of informed consent by the University of Cincinnati Institutional Review Board.

Clinical data was obtained from the electronic health record, including essential demographic information, injury characteristics, and duration of clinical monitoring. Determination of consciousness was made through documentation of command following confirmed by GCS motor sub score of 6 documented during hourly bedside clinical assessments during the monitoring period. Patients were stratified as conscious if they did so at least 6 hours prior to stopping monitoring. Those who did not follow commands by the end of the monitoring period were considered to be unconscious.

Perfusion was measured using thermal diffusion flowmetry (TDF) (Hemedex, Inc; Waltham, MA). EEG was recorded using a Natus EEG system (Natus Medical Incorporated; Middleton, WI). EEG data was sampled at 256 Hz and perfusion measurements were recorded at 1Hz. EEG data was analyzed from the F4 electrode when the TDF probe was placed on the right hemisphere or F3 when the TDF probe was placed on the left hemisphere. MNM data were recorded simultaneously using the bedside Moberg Component Neuromonitoring (CNS) system (Natus Medical Incorporated; Middleton, WI).

### 2.3 Non-invasive systems

Two non-invasive diffuse correlation spectroscopy (DCS) instruments, one time-gated (1064 nm)[49,50] and one continuous-wave (785 nm)[51], were placed on the scalp near EEG electrodes to monitor cerebral blood flow. The optical blood-flow indices they produced were recorded in parallel with an invasive thermal-diffusion flowmetry probe and showed close temporal agreement.

### 2.4 Data segmentation and labeling

All data were analyzed in MATLAB (MathWorks Inc). Both EEG and perfusion data were first divided into separate segments due to the TDF probe recalibration to ensure accurate perfusion data. Segments that were within the first six hours and last six hours of measurement duration were considered to balance the data between subjects and to better reflect the conscious and unconscious states. To ensure accurate perfusion data was used, only the first thirty minutes after probe recalibration from each segment were used for analysis. The EEG data was then bandpass filtered from 0.5 to 30 Hz using EEGLAB [1] to remove signal drift and noise. After filtering, the EEG power spectral density was calculated using Welch’s method at one-second intervals with a 30-second window to obtain a value every 1 Hz to match the sampling rate of the perfusion data. The power spectral density was then used to calculate the power in the four frequency bands, alpha (8-12 Hz), beta (12-30 Hz), delta (1-4 Hz), and theta (4-8 Hz), as well as the total power in the 1-12 Hz range [11,12]. Since 12-30 Hz band was too noisy, up to 12 Hz was considered in total power. Alpha / Delta, referred to ADR, and Alpha / (Delta + Theta), denoted as ADTR, ratios were then calculated. Perfusion data was also divided into separate frequency bands using Welch’s method to assess low frequency oscillations. Specifically, band IV (0.027-0.073 Hz) and band V (0.01-0.027 Hz) which correspond to neurogenic and endothelial contributions to cerebral autoregulation [52,53]. Additionally total frequency power from 0 to 0.5 Hz defined as band All was determined for the perfusion data. Since mean arterial pressure was not measured, the normalized frequency bands of the perfusion data were analyzed as a surrogate for cerebral autoregulation. Wavelet coherence analysis was performed between ADR and ADTR with perfusion defined as ADR-WCOH and ADTR-WCOH respectively using the same process as described in reference [54]. After the computation of the EEG frequency bands and wavelet coherence, each parameter from the 30-minute segment was averaged to a single value for that segment for machine learning (ML) analysis. A simple flow chart of the analysis is shown in Figure 2.

**Figure 2.**
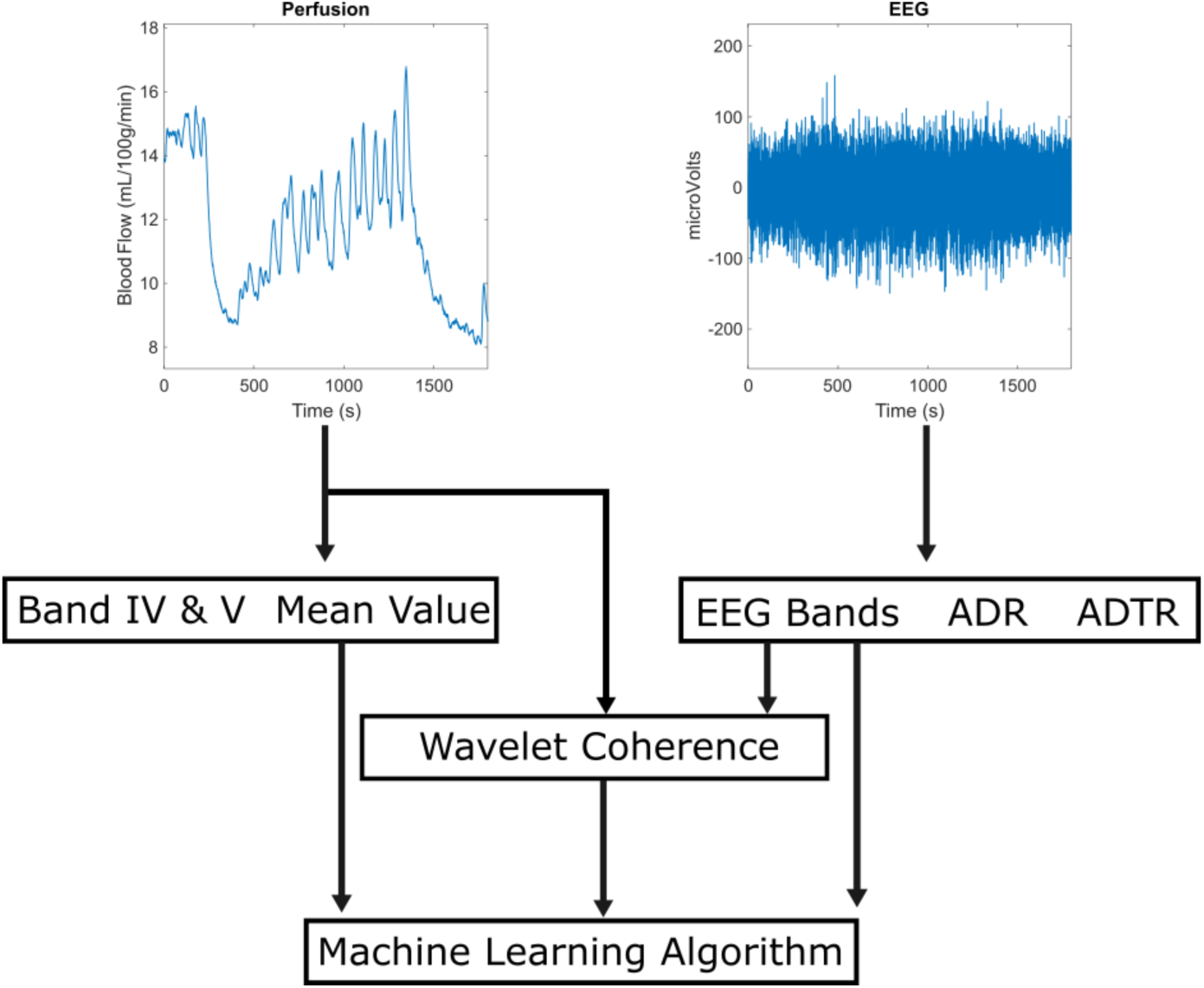
The flowchart of the data processing.

### 2.5 Preprocessing

The raw dataset (144 segments) was then normalized using Eq. (1), to ensure that the data features have unit-lengths, preserving the data structure and improving the accuracy of subsequent processes.

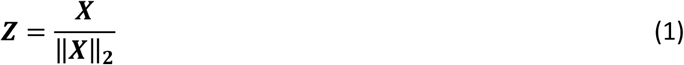

In the equation above, **𝑍**represents the standardized value and **𝑋**stands for the original value.

To address the issue of data imbalance, the data was weighted. Initially, the dataset comprised 10 conscious patients (29 segments) and 26 unconscious patients (115 segments), and through weighting the conscious group was weighted more to ensure fair classification.

### 2.6 Model selection

The entire dataset was randomly shuffled to avoid any bias. This study employed a K-Fold cross-validation technique to validate and assess performance. Since the aim of the models is to identify the binary consciousness state of each segment, K-Fold cross-validation is not applied on patient level without any data leakage, and a separate set of patients (non-invasive patients) are considered for final tests. Essentially, the data is split into K folds, using a fold to train the model, and the remaining fold to test the model. The K-Fold method evaluated and optimized hyperparameters of K-nearest neighbors (KNN), random forest (RF), linear regression (LR), gradient boosting (GB), gaussian naive Bayes (GNB), and neural network (NN) models before testing on the remaining data. Modeling was performed utilizing the Scikit-Learn and SALib libraries [55–57].

ADTR, ADR, TotPower, band IV, band V, band All, ADTR-WCOH, and ADR-WCOH were used as input features for each of the models. TotPower refers to the total EEG power from the 1-12 Hz range.

### 2.7 Details of the ML models and metrics

A detailed overview of the ML models used, along with their key parameters, is provided in Table 1. The models include KNN, RF, LR, GB, GNB, and NN. Each model’s description encompasses the primary parameters set for further experiments, ensuring a comprehensive evaluation of their performance. These parameters were optimized based on a random grid-search integrated with the K-Fold method [58–60]. Table 1 encapsulates the vital aspects of each model, highlighting the thorough optimization and selection process undertaken in this study.

**Table 1.** Summary of machine learning models and their tuned hyperparameters.

| Model | Description |
| --- | --- |
| KNN | Number of neighbors: 3, algorithm: auto, leaf size: 30, distance metric: Euclidean |
| RF | Number of trees: 400, maximum depth: 12, minimum samples to split: 6, minimum samples per leaf: 3, features considered per split: 20%, bootstrap: True, out-of-bag score: True, random state: 42 |
| LR | Penalty: L2 regularization, solver: LBFGS, maximum iterations: 100, regularization strength: 1.0, fit intercept: True |
| GB | Number of boosting stages: 100, learning rate: 1.0, maximum depth: 1, random state: 42, loss function: deviance, criterion: Friedman MSE |
| GNB | Variance smoothing: 1e-9 |
| NN | Hidden layer sizes: 100 neurons, maximum iterations: 300, random state: 42, activation function: ReLU, solver: Adam, regularization term: 0.0001 |
| LBFGS= Limited-memory Broyden–Fletcher–Goldfarb–Shanno; ReLU= Rectified Linear Unit. MSE= Mean Squared Error |  |

The study employed several performance metrics to evaluate the ML models. These metrics include accuracy and receiver operating characteristic area under the curve (ROC-AUC). Accuracy measures the proportion of correctly classified instances out of the total instances. ROC-AUC provides a measure of the model’s ability to distinguish between classes. On top of these metrics, a confusion matrix offers a more granular view of the model’s performance. It provides detailed insights into the true positives, true negatives, false positives, and false negatives. This level of detail is crucial for understanding how well the model distinguishes between different classes and where it may be making errors. By examining the confusion matrix, one can gain a deeper understanding of the model’s strengths and weaknesses, leading to more informed decisions about the selection of the best model.

### 2.8 Outcome Assessment

Ground truth labels for classification were derived from clinical documentation of command-following ability during the EEG/CBF monitoring window. Patients who demonstrated consistent, reproducible command following were labeled “conscious”, while those without any such behavioral evidence were labeled “unconscious”. CRS-R scores and clinical assessments were used to support classification where available.

### 2.9 Statistical Analysis

Classification performance was evaluated using standard metrics including accuracy and area under the receiver operating characteristic curve (ROC-AUC). Additional metrics such as precision and F1 score were calculated where appropriate.

## 3 Results

### 3.1 Demography and Statistical analysis of the dataset

The study included 35 patients (mean age: 35.5 years old, range: 18-85), predominantly male (91.4%). The mean initial GCS score was 6.3, with 37.1% undergoing craniectomy prior to intracranial pressure monitoring. Most bolt placements were right-sided (68.6%). The average ICU and hospital lengths of stay were 12.9 and 16.3 days, respectively. Glasgow Outcome Scale (GOS) scores ranged from 1 to 5, with 28.6% (n=10) achieving a good recovery (GOS 5) and 22.9% (n=8) exhibiting moderate disability (GOS 4).

The first patient monitored noninvasively was a male in his 20s. The second was a male in his 40s. The first patient exhibited severe traumatic brain injury with a reduced level of consciousness (GCS 7T) and radiological evidence of a left subdural hematoma with midline shift. The second patient displayed severe traumatic brain injury characterized by extensor posturing, an unresponsive state (GCS 3T), and imaging showing an 8 mm subdural hematoma with midline shift.

A detailed statistical summary of the features is provided in Table 2. These features are ADTR, ADR, TotPower, Band IV, Band V, Band All, ADTR-WCOH, and ADR-WCOH. Preliminary analysis compared EEG and CBF-derived features between unconscious and conscious groups. The conscious group exhibited higher mean TotPower (TotPower: 0.58 vs. 0.45), while the nonconscious group showed elevated mean ratio-based features (ADR: 0.39 vs. 0.02; ADTR: 0.0.23 vs. 0.05), suggesting moderate effects.

**Table 2.** Summary of statistical parameters.

| Group | Statistics | TotPower | ADR | ADTR | WCOH<br>ADR | WCOH<br>ADTR | BandV | BandIV | BandAll |
| --- | --- | --- | --- | --- | --- | --- | --- | --- | --- |
| 0* | Mean | 0.58 | 0.06 | 0.05 | 0.02 | 0.02 | 0.003 | 0.008 | 0.028 |
|  | Std Dev | 0.19 | 0.05 | 0.04 | 0.02 | 0.02 | 0.001 | 0.005 | 0.016 |
|  | Min | 0.08 | 0.01 | 0.01 | 0.00 | 0.00 | 0.001 | 0.001 | 0.004 |
|  | Q1 | 0.46 | 0.01 | 0.01 | 0.01 | 0.01 | 0.002 | 0.004 | 0.015 |
|  | Median | 0.56 | 0.03 | 0.03 | 0.01 | 0.01 | 0.003 | 0.006 | 0.029 |
|  | Q3 | 0.74 | 0.09 | 0.07 | 0.04 | 0.04 | 0.004 | 0.012 | 0.038 |
|  | Max | 0.89 | 0.20 | 0.17 | 0.06 | 0.06 | 0.005 | 0.018 | 0.069 |
| 1* | Mean | 0.45 | 0.39 | 0.23 | 0.01 | 0.01 | 0.002 | 0.005 | 0.019 |
|  | Std Dev | 0.22 | 0.87 | 0.39 | 0.01 | 0.02 | 0.002 | 0.004 | 0.015 |
|  | Min | 0.02 | 0.00 | 0.00 | 0.00 | 0.00 | 0.000 | 0.000 | 0.001 |
|  | Q1 | 0.29 | 0.05 | 0.04 | 0.00 | 0.00 | 0.001 | 0.001 | 0.008 |
|  | Median | 0.47 | 0.12 | 0.09 | 0.01 | 0.01 | 0.001 | 0.003 | 0.015 |
|  | Q3 | 0.60 | 0.29 | 0.22 | 0.02 | 0.02 | 0.003 | 0.007 | 0.023 |
|  | Max | 0.91 | 6.32 | 2.17 | 0.07 | 0.07 | 0.007 | 0.020 | 0.064 |

\* 0 means conscious while 1 refers to unconscious.

### 3.2 Correlation analysis

The correlation factors, which depict the relationships between various features and outcome, is shown in Figure 3. The features show no linear correlation within the feature domain (correlation < 1). When combining features, those with high collinearities (>0.8) are not used at the same time. For example, ADR and ADTR are never used together.

**Figure 3.**
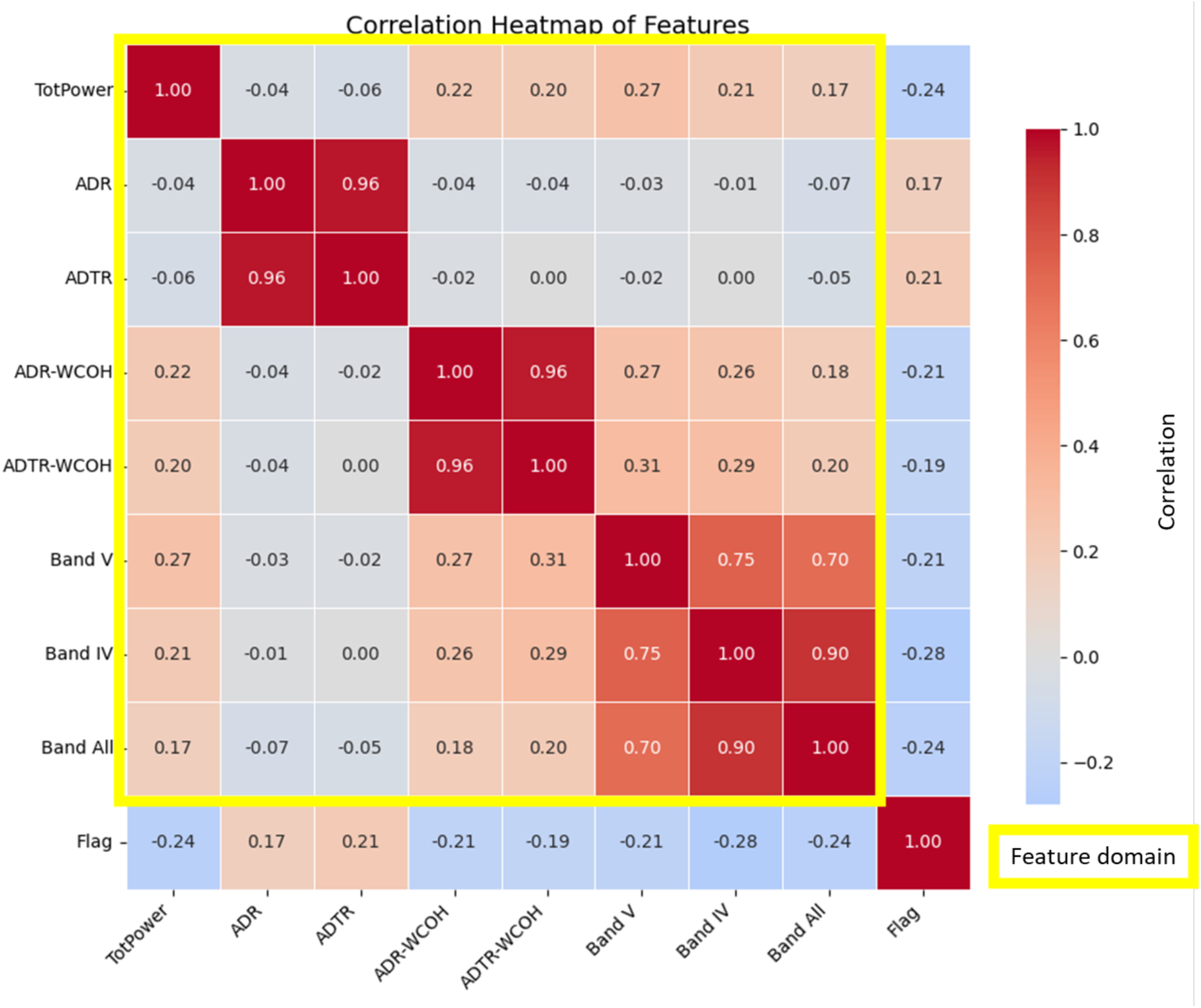
Correlation factors: Features with high correlation (>0.8) should not be used together.

### 3.3 Machine Learning model classification and interpretation

First, we tested all valid combinations (see Figure 3 for correlation factors) of ADTR, ADR, TotPower, Band IV, Band V, Band All, ADTR-WCOH, and ADR-WCOH for all models with RF, which provided the best results overall. Results from all the models are shown in supplemental data (Table S1). Table 3 summarizes the performance metrics of the RF model evaluated using 5-fold cross-validation of data. Note that correlated features (see Figure 3) are not considered together. This analysis reveals that the [ADR, TotPower, Band V] combination most consistently achieved the highest scores, closely (0.01 difference) followed by the [ADR, TotPower, Band All] and [ADR, TotPower, Band IV] combinations. As observed in Table 3, WCOH-ADR benefits most from addition of perfusion features (61-69 %), but its best ROC-AUC (0.68) and accuracy (0.76) remain lower than the others. More metrics (e.g., balanced accuracy, F1 score, Area Under the Precision-Recall Curve (AU-PR), ROC-AUC- 95% Confidence Intervals (CIs), etc.) are provided in supplemental data (Table S2) for the top combinations.

**Table 3.**
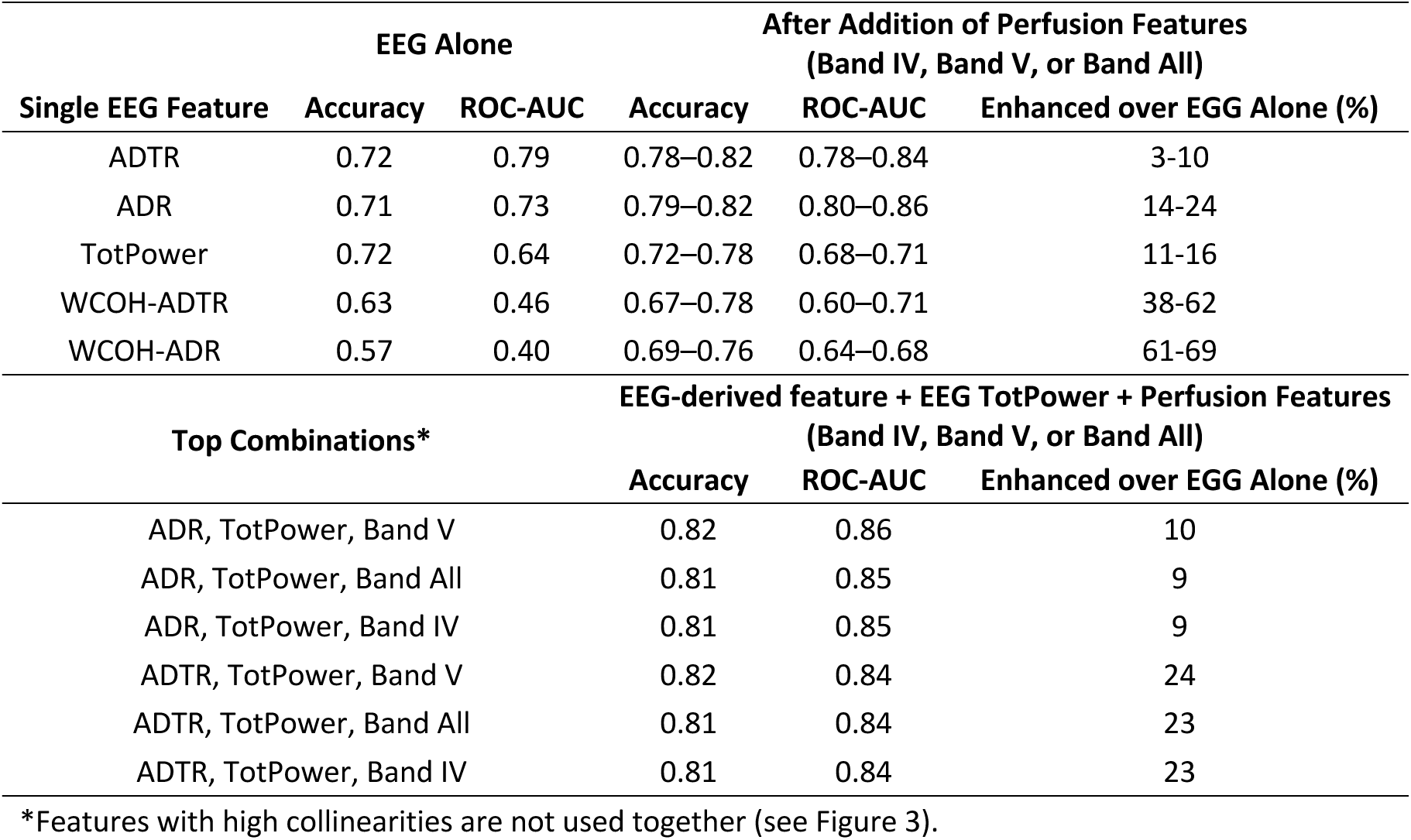
Effect of adding perfusion features to EEG on machine-learning performance (measured by 5-fold cross-validation).

Figure 4 shows the confusion matrices of the best combination of features (ADR, TotPower, Band V) for various ML models averaged on 5 folds. Overall, the RF (Figure 4 (f)) outperformed all others, misclassifying only 5.2 samples in total. The LR (Figure 4 (c)) offered the most balanced results, correctly predicting 69 % of observations in each class, but at the cost of a high overall error count (9 misclassifications).

**Figure 4.**
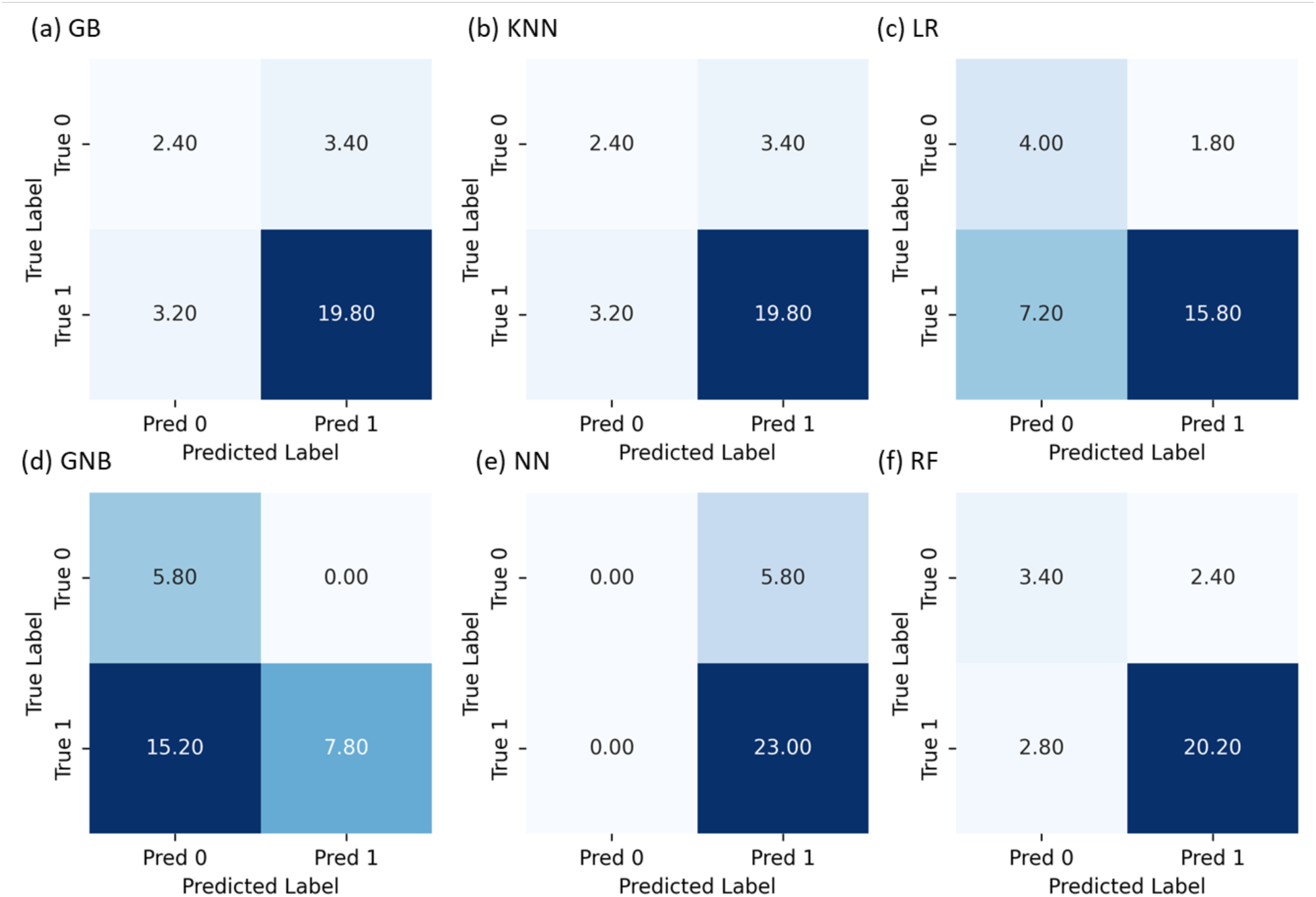
Mean confusion matrices for the optimal feature set (ADR, TotPower, Band V) on the full dataset. True condition (conscious = 0 / unconscious = 1) is plotted on the y-axis and predicted condition on the x-axis, with panels showing: (a) GB, (b) KNN, (c) LR, (d) GNB, (e) NN, and (f) RF models.

### 3.4 Non-invasive clinical results

Figure 5 shows (a) Late Gate (LG) DCS, (b) Early Gate (EG) DCS, and (c) CW-DCS clinical measurements. Overall, there are good correlations between non-invasive and invasive counterparts. The gap observed between approximately 3500 and 4000 seconds in time-gated data is a consequence of an automatic calibration cycle implemented by the thermal diffusion flowmetry (TDF) device. When a substantial fluctuation in cerebral blood flow occurs, often triggered by interventions such as a neurological examination or adjustments in sedation, the TDF device initiates a recalibration process to ensure the accuracy and reliability of subsequent measurements. During this recalibration, data acquisition is temporarily suspended, leading to the brief gap in the recorded signal. This behavior is inherent to the clinical operation of the device and is essential for maintaining measurement fidelity in dynamic physiological conditions.

**Figure 5.**
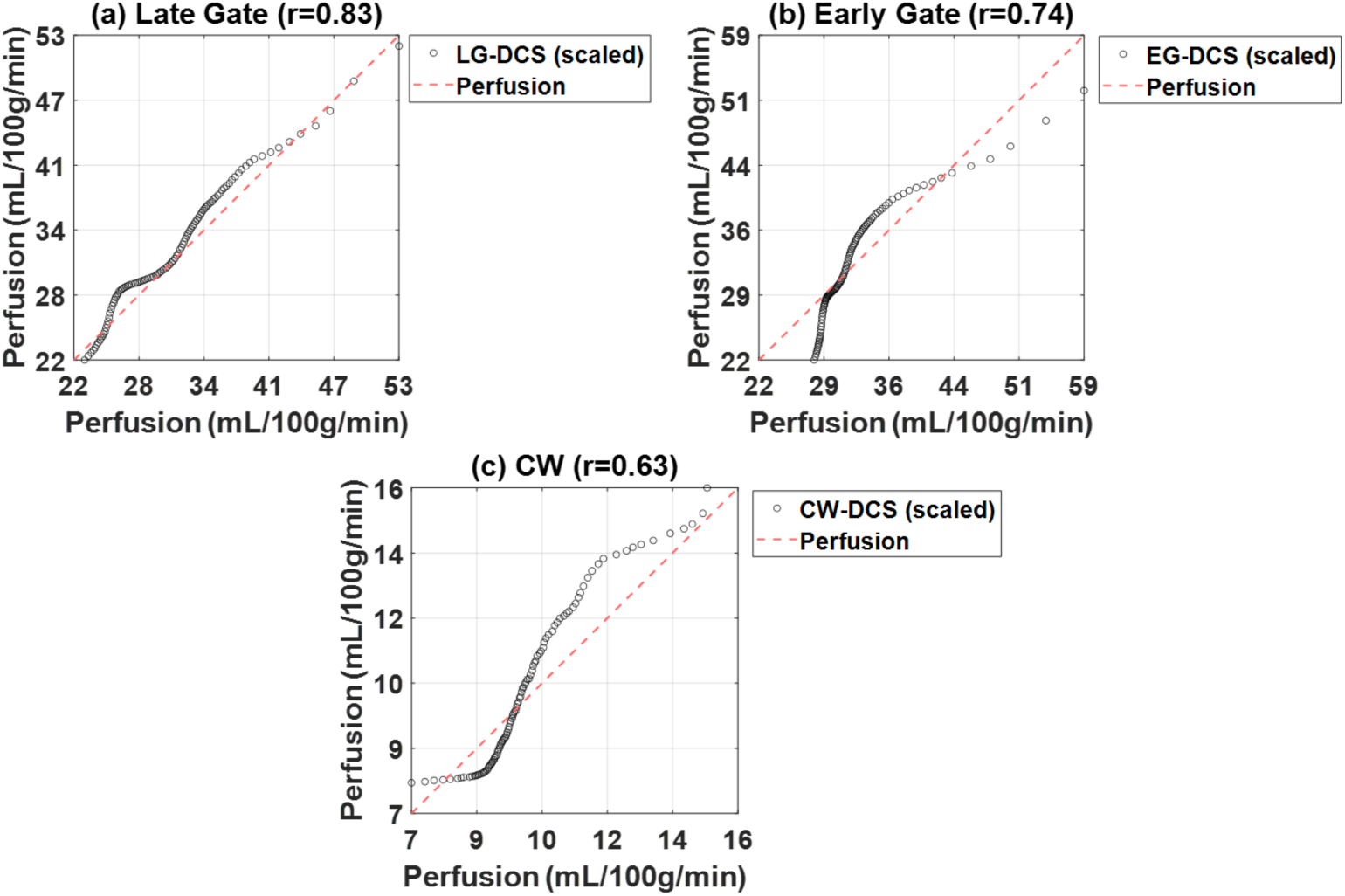
Noninvasive data captured using (a) LG-DCS, (b) EG-DCS, and (c) CW-DCS.

### 3.5 Non-invasive ML prediction

The RF was further evaluated with the test data from the noninvasive methods of CBF (i.e., LG-DCS, EG-DCS, and CW-DCS) and their invasive counterparts. As shown in Figure 6, the RF accurately predicted all instances of both invasive and noninvasive measures of CBF of the final test data.

**Figure 6.**
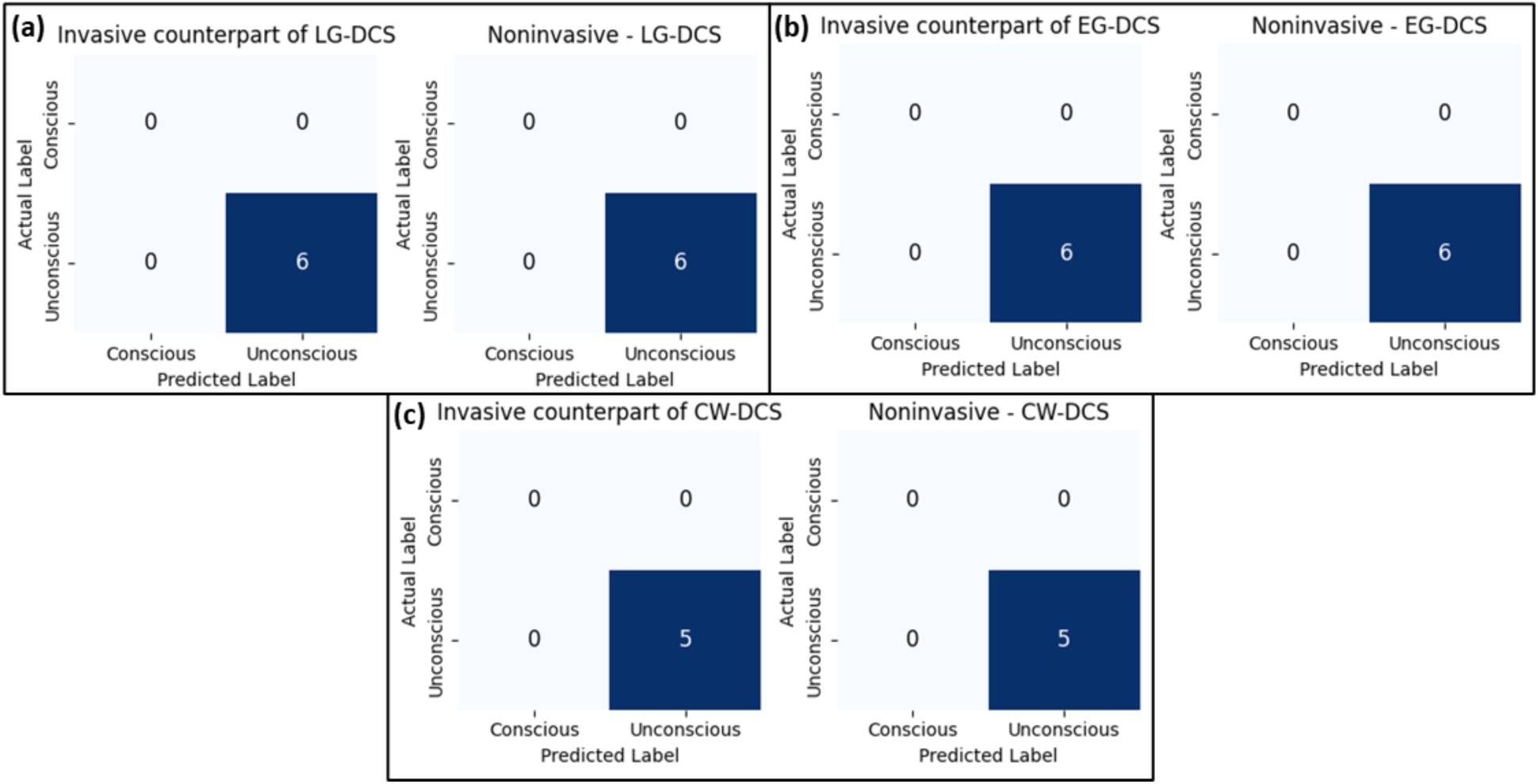
The best model’s (RF) confusion matrices for the best combination of features (ADR, TotPower, Band V) between invasive and non-invasive methods for (b) LG-DCS, (b) EG-DCS, and (c) CW-DCS.

## 4 Discussions and conclusions

Accurate classification of consciousness levels in patients is essential for effective clinical care and prognosis. Evaluating neurological status and predicting outcomes for comatose patients is difficult due to clinical status and treatment plans, which can be improved by using machine learning to combine measures of cerebral perfusion and cortical activity. Results show that combing both EEG and cerebral perfusion parameters performed better than just EEG parameters only.

### 4.1 Novelty

Due to the complexity and size of data that comes from MNM, many studies have been focused on developing ML models to help identify DOC [61–66]. Table 4 summarizes additional studies on ML modeling of consciousness status across diverse patient populations, further illustrating the transformative potential of these technologies in clinical practice. Collectively, these studies demonstrate the growing impact of ML and advanced neuroimaging in improving the diagnosis, prognosis, and management of DOC and TBI, paving the way for more personalized and effective patient care.

**Table 4.**
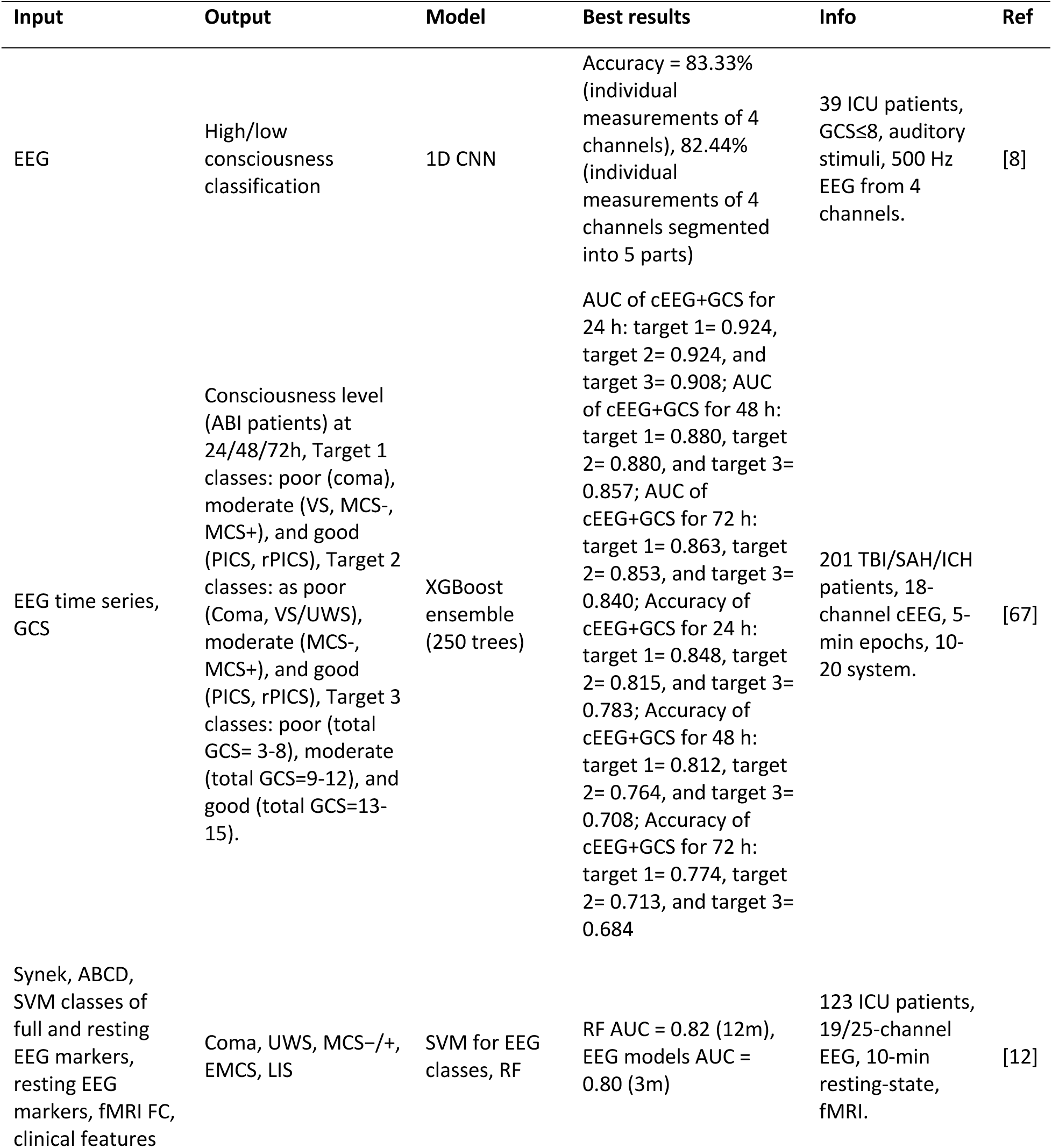
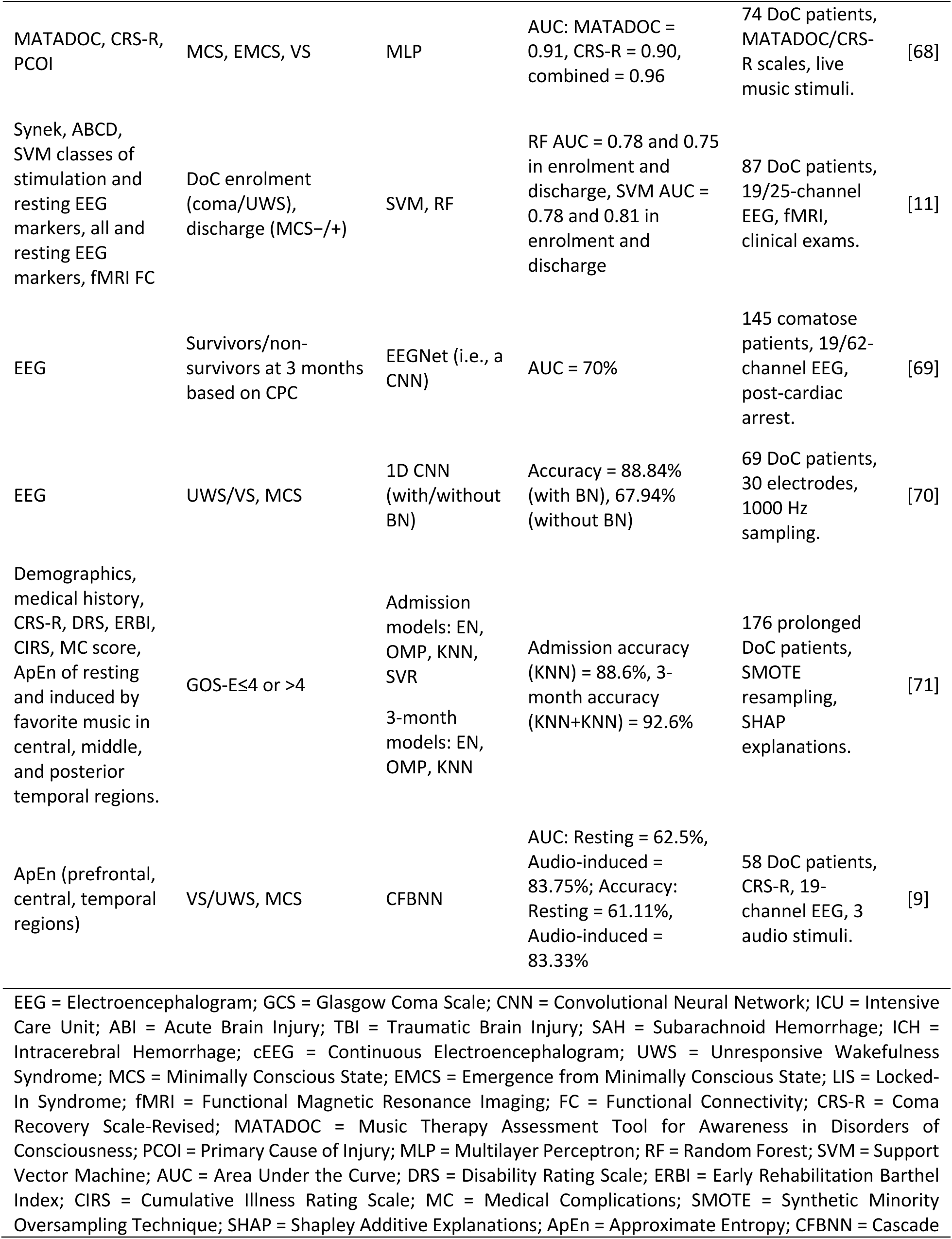
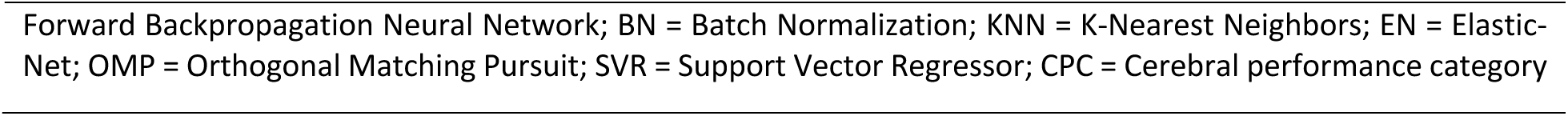
Summary of previous studies.

Most studies are too complex to replicate concerning either the proposed ML models or input and output features. This study introduces novel models for classification of consciousness by examining possible combination of physiologically interpretable features including ADR, TotPower, Band IV, Band V, Band All, ADTR-WCOH, and ADR-WCOH. Previous studies have utilized fMRI and EEG, in which fMRI encodes information about brain activity and functional connectivity [11,12,72]. However, fMRI is more expensive and does not provide bedside measurement over time compared to perfusion (CBF) [73]. Unlike previous studies that rely on complex or high-dimensional feature sets, our method focuses on a simplified feature domain to provide a concise picture of the neurovascular unit. By integrating these features into 6 simple and easily reproducible models, we aim to achieve robustness measured by performance metrics.

### 4.2 Data Preprocessing and Model Selection

The preprocessing steps, including standardization and weighting, were crucial in ensuring the robustness and accuracy of the ML models. By weighting the dataset, we mitigated the risk of bias and improved the generalizability of the models. The use of K-Fold cross-validation further enhanced the reliability of our findings by preventing overfitting and underfitting, which are common challenges in ML, especially with smaller datasets.

### 4.3 Model Performance

Among the evaluated models, the RF model emerged as the top performer in the K-Fold cross-validation. The confusion matrices highlighted RF as the top model, with the minimum total misclassifications. The superior performance of RF can be attributed to its ability to handle the variability in the dataset effectively [74].

### 4.4 Effect of the Perfusion Features

The integration of perfusion features (Band IV, Band V, or Band All) with EEG-derived metrics significantly enhances the classification of conscious and nonconscious states up to 69%, as demonstrated by systematic improvements in accuracy and ROC-AUC (Table 3). Universally, the top-performing combination of parameters included ADR/ADTR, TotPower, and one of the perfusion bands, with [ADTR, TotPower, Band V] achieving the best classification results (ROC-AUC: 0.86, accuracy: 0.82). This underscores the synergistic role of perfusion Band V’s hemodynamic fluctuations in amplifying EEG-derived amplitude (ADR/ADTR) and spectral power (TotPower). Combinations incorporating band All or IV also performed closely (i.e., [ADTR, TotPower, Band IV or All]: ROC-AUC 0.85). This is expected as band V has a relatively large collinearities with band IV and all (respectively, 0.75 and 0.70 in Figure 3), leading to a similar effect. The effectiveness of band V suggests low-frequency perfusion signals are physiologically pivotal for consciousness discrimination. Notably, even wavelet coherence features (ADTR-WCOH and ADR-WCOH), which showed limited utility (standalone ROC-AUC: 0.46 and 040., respectively), achieved marked gains when fused with multiple features (e.g., [ADTR-WCOH, TotPower, Band V] with ROC-AUC of 0.71 and [ADR-WCOH, TotPower, Band V] with ROC-AUC = 0.68 in Table S3). These results highlight that optimal classification hinges not on isolated features but on multimodal frameworks leveraging neurovascular coupling.

### 4.5 Noninvasive Performance

The study demonstrates that the RF model functions effectively regardless of whether the input data is invasive or non-invasive. By applying the same algorithm to the different measures of CBF from DCS along with traditional invasive counterpart measurements, the resulting predictions show consistency. This outcome highlights the robustness of the RF, suggesting that non-invasive methods can provide reliable insights into cerebral blood flow equivalent to those obtained through invasive techniques. This is while non-invasive methods offer clear advantages in terms of patient safety and comfort.

### 4.6 Implications and Future Directions

The study’s results have important implications for assessing the binary consciousness state in patients. The high accuracy and reliability of the RF model indicate its potential for clinical use, providing healthcare professionals with a reliable tool for patient evaluation. Future research should aim to expand the dataset to include a more diverse patient population and explore the integration of additional features to enhance model performance further. Additionally, developing user-friendly interfaces for ML models could facilitate their adoption in clinical settings, enabling real-time monitoring and decision-making. Incorporating advanced techniques, such as transformers and ensemble methods, may also offer further improvements in classification accuracy and robustness.

Moreover, this study aimed at the general classification of comatose patients into conscious and unconscious groups. A future extension will cover a detailed classification of consciousness into different levels. Additionally, this study did not use any evoked potentials (e.g., auditory [75]) to assess cortical activity. For instance, mismatch negativity (MMN) might be used, which is an event-related potential (ERP) component that occurs in response to deviant stimuli [76]. It reflects the brain’s automatic detection of deviations from a regular pattern, even without conscious attention. MMN is valuable in clinical research for assessing auditory discrimination abilities and has been used in comatose prediction [77,78].

### 4.7 Limitations

Despite the promising results, the study has some limitations. The relatively small sample size may limit the generalizability of the findings. Future studies should aim to address these limitations by increasing the sample size and exploring alternative methods for handling data imbalance.

Another limitation of the study is the use of an invasive device to monitor cerebral perfusion. Depending on the patient’s status, invasive measures of perfusion are not feasible thus an alternative is required. Traditional neuroimaging techniques like magnetic resonance imaging (MRI) and functional-MRI (fMRI) face significant limitations. They are expensive, non-portable, and unsuitable for patients with metallic implants or frequent monitoring [79,80] . Alternatively, diffuse optical techniques such as functional near-infrared spectroscopy (fNIRS) and Diffuse Correlation Spectroscopy (DCS) offers portability and bedside usability [81,82]. fNIRS has demonstrated the ability to detect covert consciousness through hemodynamic responses to stimuli (e.g., familiar music) and task-based paradigms like motor imagery [21,25]. Studies show that fNIRS can identify resting-state networks and command-driven activity in behaviorally unresponsive patients, with low-frequency oscillations (LFO) in oxyhemoglobin signals correlating with consciousness levels [25] . However, fNIRS suffers from limited spatial resolution, sensitivity to superficial tissue contamination [83], and challenges in distinguishing brain-specific signals [84] . Diffuse correlation spectroscopy (DCS) is an optical method that noninvasively measures cerebral blood flow and has been used in previous studies to monitor critically ill patients [49–51,85–89] . This method has also been validated against other blood flow monitoring methods [90–92] thus showing its reliability and will be used in the future as an alternative to invasive monitoring. More recently, impaired neurovascular coupling was observed in patients undergoing extracorporeal membrane oxygenation (ECMO), with encephalopathic (non-command-following) patients showing significantly weaker correlations between EEG and cerebral blood flow (measured by diffuse correlation spectroscopy) compared to awake patients [86]. Lastly, mean arterial pressure was not analyzed with respect to cerebral autoregulation. Thus, only the normalized frequency spectra of the perfusion data were used to evaluate cerebral autoregulation. In the future, blood pressure will be recorded and correlated with perfusion data to give a better estimation of cerebral autoregulation.

### 4.8 Conclusions

Accurate consciousness assessment in neuro-intensive care is critical for timely intervention. Our machine learning framework, integrating EEG and cerebral blood flow (perfusion) metrics, achieves robust classification (ROC-AUC: 86%) using a random forest model, outperforming EEG-only approaches by up to 69%. By capturing complementary neurovascular signatures, this multimodality enhances clinical decision-making for treatment and rehabilitation planning.

## Code and Data Availability

Data may be provided by the corresponding author upon reasonable request.

## Data Availability

Data may be provided by the corresponding author upon reasonable request.

## Acknowledgements

The authors acknowledge the funding support from NIH R01 (NIBIB Brain Initiative (7R01EB031759-03). ChatGPT (GPT-4) was used to assist with grammar and language editing during the preparation of this manuscript.

